# Globally Coherent Weekly Periodicity in the Covid-19 Pandemic

**DOI:** 10.1101/2020.06.02.20120238

**Authors:** C. S. Unnikrishnan

## Abstract

After the major exponential phase, the Covid-19 pandemic is on a milder rate of increase globally, and past the peak in many countries. In this phase a marked systematic weekly periodicity in the Covid-19 attack has become clear, the study of which could be useful for the long term strategy to deal with the virus and the pandemic. The most important aspect of this strong weekly modulation is its global nature, phase-coherent over most of globe, independent of the geographical location. The fact that the same periodicity is now evident even those countries Asia and Africa, where it was absent or not prominent earlier, suggests the possibility that the pattern could be an early indicator for significant community transmission. The global coherent periodicity with a time scale that agrees with the time scales of virus incubation and infection may be important in tracking the long term interaction of the virus with the host.

## The Global Data on the Covid-19 Pandemic

The Covid-19 virus attack is the first global pandemic in a century. It has changed drastically the means and styles of life. Several studies on the pathological aspects of the virus, and on the possible cures and prevention are rapidly progressing. However, at this juncture, there is much uncertainty about the time required to return to near normalcy of life. There is a large amount of data accumulated by now, due to the global nature of the pandemic. It is clear that any stable systematic feature observed in the global data is potentially very useful in handling the pandemic. It is with this view that I point out a marked weekly modulation, easily noticeable in the daily data over the past several weeks, in the daily reported cases as well as in the daily deaths due to Covid-19. The depth of modulation in the global data is as large as 30%, with enormous statistical significance. Whether this has to do with any characteristic of the virus or in the virus-host (human) interaction can only be answered after focussed studies on the biological and medical aspects. It is also possible that some extraneous sociological factor in reporting and documenting can introduce spurious periodicity in pandemic data, though this possibility is of a smaller chance, as suggested by the strength and coherence of the modulation. While a significant correlation between the daily reporting of cases and the number of daily deaths could be expected, the root cause of the stable and globally universal modulation in the number of cases reported daily needs close attention. As I will discuss, while this periodic pattern was strongly manifest in the data from the USA, South America, and Europe since April 2020, it was absent earlier in Russia, in the countries in Asia and in most of Africa, where are rate of increase in the pandemic was relatively slow and rate of death was smaller. However, the same pattern is now visible in some of this countries, like India. This suggest that the pattern could be an indicator for the stage of community transmission of Covid-19.

The data collected from various national sources of the countries are tabulated meticulously by many sources, and it is available as open source information [1, 2, 3]. As an indicative, yet impressive pointer, I show the 3-day running average of the daily deaths reported in the USA, Europe, South America (Brazil), Asia, and Africa in a single chart, along with the world aggregate (figure 1). The data, which is based on the compilation by the European Centre for Disease Prevention and Control, and the plot is from the site “Our World in Data” [2]. What is unmistakable is the weekly modulation that is coherent over 3 separate continents (North America, South America and Europe). The data from Asia, where the number of deaths have been relatively low, is yet to show an unambiguous trend, but there are some indication in the data on the daily confirmed cases (figure 3). In any case, the stable and well defined weekly periodicity in the global data is indicative of a significant clue about the Covid-19 pandemic.

**Figure 1:**
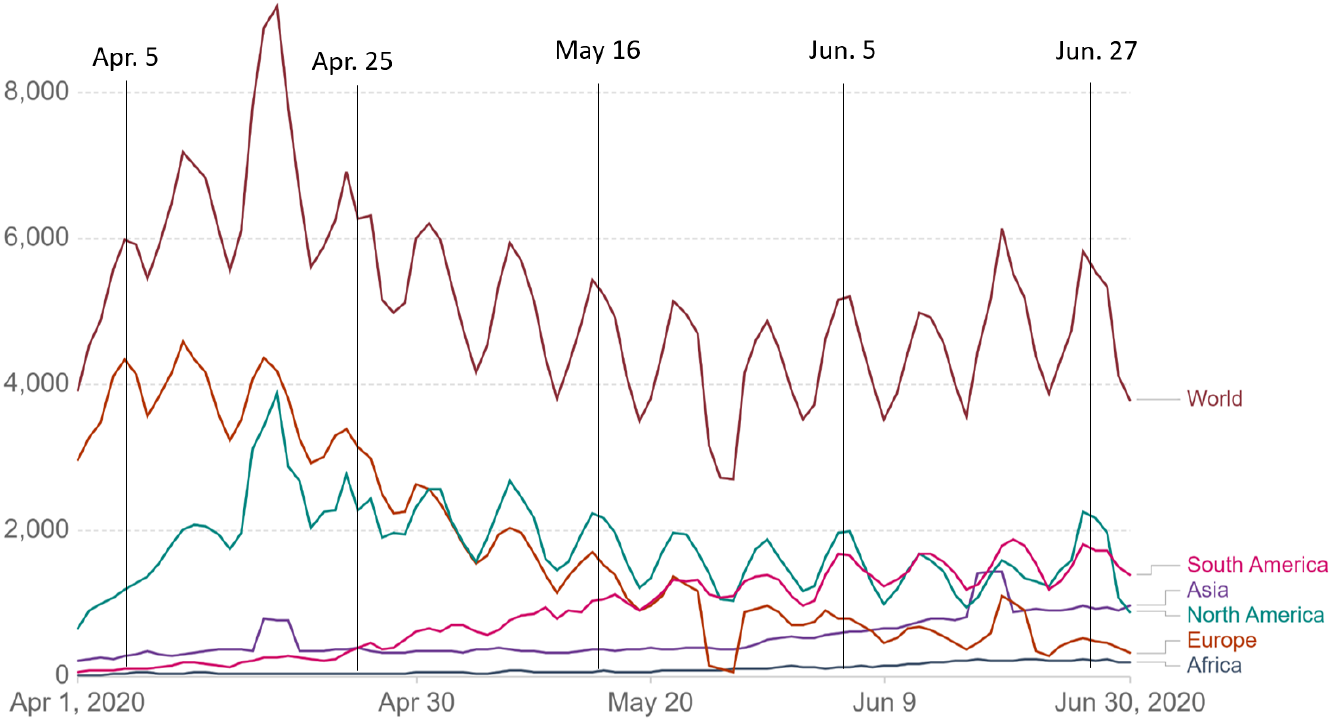
The number of daily Covid-19 deaths, plotted as 3-day running average. Left: for the USA, Europe, South America, Asia and Africa, along with the world aggregate. The strong weekly modulation with remarkable coherence all over the globe is evident. (all graphs were generated with the compiled data and the grapher tool at the site ourworldindata.org).

**Figure 2:**
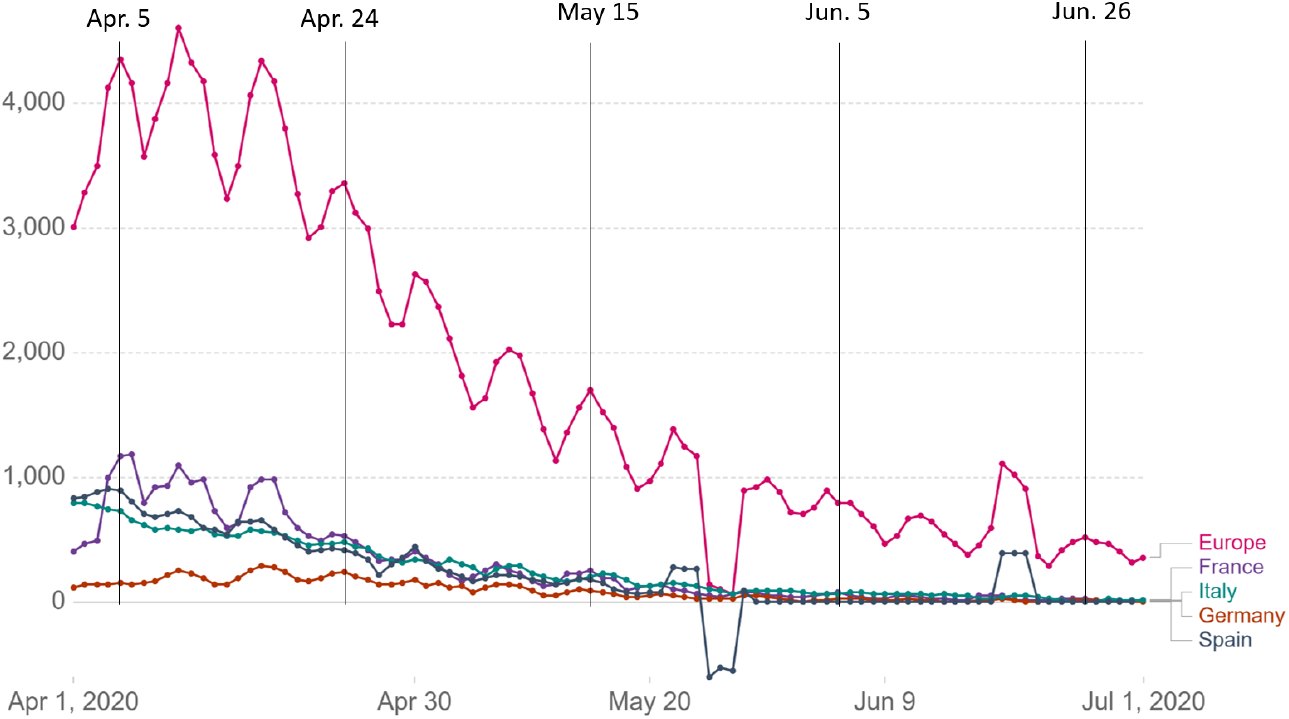
Daily Covid deaths in the countries Italy, France, Germany, Spain, and the UK, along with the European aggregate. The coherence of the weekly modulation is strongly evidenced in the aggregate.

**Figure 3:**
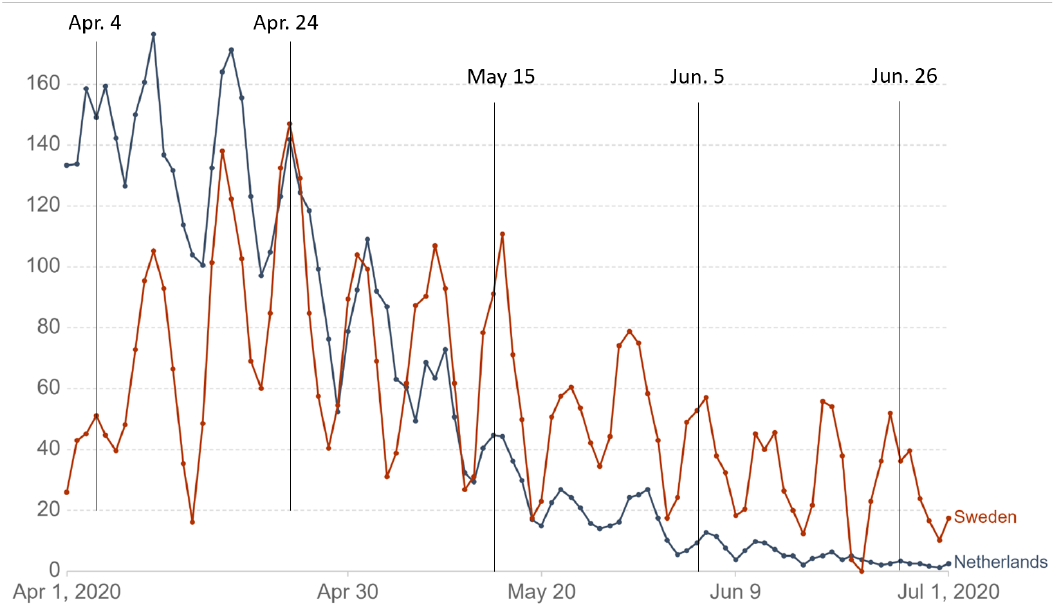
Daily death rates in Sweden and Netherlands, showing the large weekly modulations. The modulation persisted even when the death rate diminished significantly.

The figure 2 shows the daily deaths in Italy, France, Germany and Spain, along with the data from the UK and the European aggregate. While the modulations in the individual traces are easily noticeable, the coherence in the modulations across the countries makes it stand up, with very high statistical significance, when the data is stacked to form a European aggregate. This is the strong evidence for some underlying and unifying reason, under-standing of which might help in dealing with the pandemic, after detailed study.

The depth of modulation varies in the European countries, but is systematics are similar, with high coherence. Even in those countries where the population and the actual number of people affected are smaller, comparatively, the same features are seen, The large depth of modulation (> 70%) in the data from Sweden is particularly noteworthy (figure 3). Sweden had allowed a lockdown-less social interaction and the pandemic is still active there, linearly rising. On the other hand, Netherlands has the situation under control, well after peak, and yet, the same weekly oscillations persist strongly. These examples are important in determining the nature of social interactions in the spreading of the pandemic.

The same global coherence and the strong weekly modulation is visible in the number of daily reported cases from everywhere in the world, where the pandemic had a strong presence so far. I show in figure 4 the daily reported cases globally, from every continent, along with the world aggregate, plotted as a 3-day running average. This global coherence is surprising, given the fact that the factors involved in the start and the spreading of the disease vary a lot between these continents. Of course, the same trend is prominently present in the data for counties in Europe as well.

**Figure 4:**
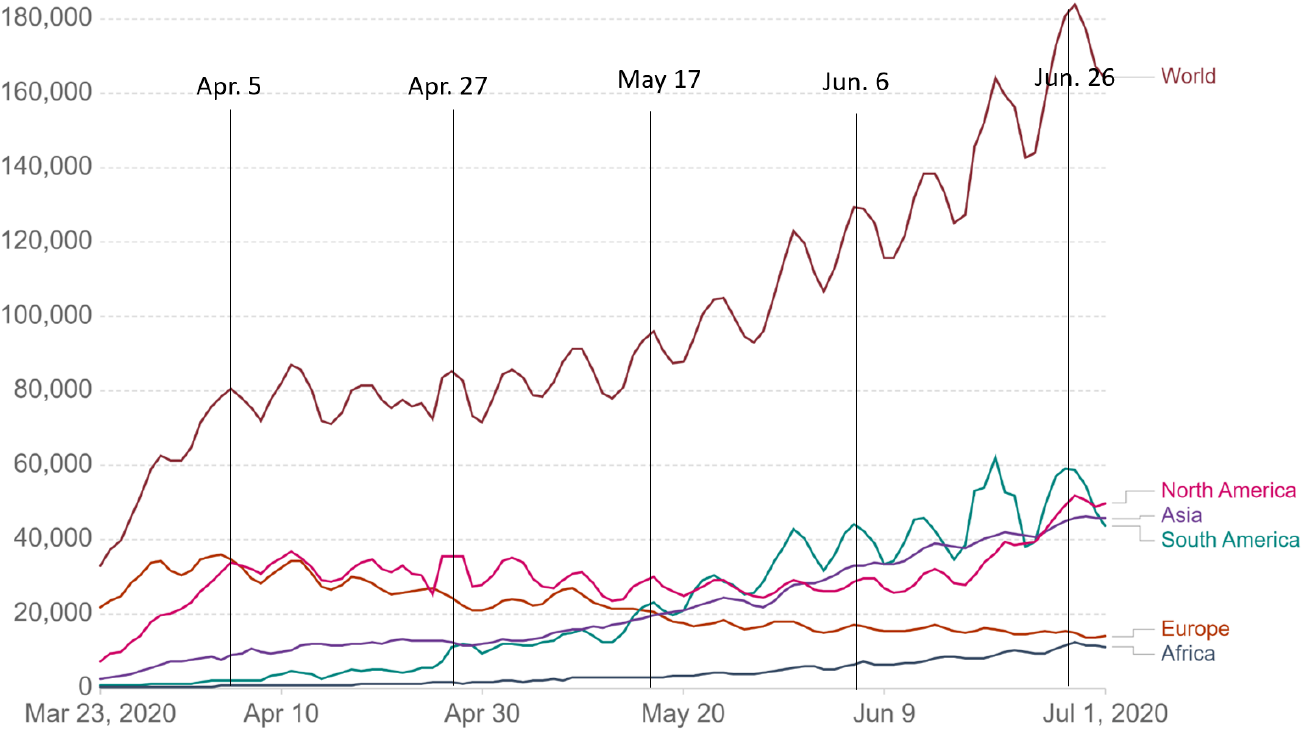
Daily confirmed Covid cases in various continents along with the world aggregate.

The weekly periodicity in the data on the daily deaths and the data on the daily reported cases is unmistakable. Of course, there could a small phase-shift between the peaks of the Covid deaths and the peaks of the daily reported cases, about 1 day (modulo7 days) but the periodicity is the same. As mentioned earlier, the depth of the modulation varies from 20% to 40%. In Asia and Africa, and also in Russia, where the Covid-19 cases and deaths have been lower, relative to the population, there was no significant evidence for a periodic weekly modulation until recently. However, as seen in the figure 5, *the situation is palpably changing, with noticeable weekly periodic pattern emerging, indicating that this is a characteristic signature in this pandemic*. If the pattern turns out to be an early indicator of community spreading of the pandemic, the modulations will certainly become stronger, as it happened in other countries in the Americas and Europe.

**Figure 5:**
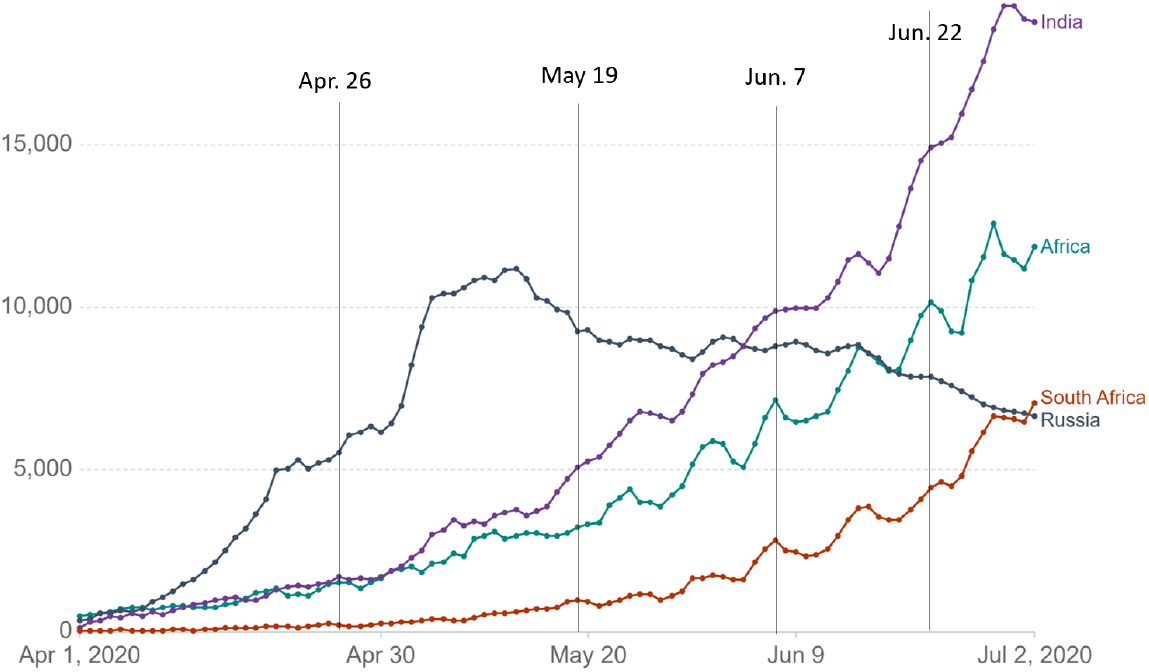
The late emerging of the weekly periodic pattern in Russia, India, and Africa, in the daily confirmed cases. A similar pattern is now discernible in the number of daily deaths as well.

## Discussion

The weekly modulation that is confirmed in both the number of daily deaths and the number of daily reported cases could be due to some temporal modulation factor in the virus itself, or more likely in the interaction of the host (humans) with the virus. Both these aspects need focussed study on the observed strong modulation with the well defined periodicity of about a week. Though the modulations remind one of the population modulations in the species interactions in ecological models [4, 5], there is no further clue available for even a speculation on that front. Self-sustaining relaxation oscillations [6], as seen in certain physical systems can also happen when a threshold is set by control strategies against the spreading of the pandemic, like a lock-down and self-imposed societal restrictions. I refrain from commenting more on the pathological aspects that might give rise to such a strong modulation. The weekly periodicity is of the same time scale as the characteristic duration of the virus-host interaction, and this may be relevant. Noticeable periodicity is decease statistics have been noted and reported previously; for example, in the cases in treatment of Tuberculosis in India, though of a much larger period (yearly) and of weak prominence [7]. However, one needs to rule out other trivial systematics arising from sociological aspects, like patterns in reporting the disease, hospital admissions etc. I tentatively rule out such causes in the Covid-19 pandemic primarily because of the global nature of the periodic modulations, evident everywhere, irrespective of the geographical location and the economic and social environment. It is perhaps important to note that in all countries, irrespective of the geographical and cultural variety and the societal patters, the number of daily reported cases as well as the number of daily deaths peaks towards the weekend and records the minima during early to mid week. This rules out simple explanations based on the patterns in reporting and administrative and logistical factors.

Whether the globally coherent periodic patterns in the Covid-19 pandemic reported here are related to some factor in the virus-host interaction or the strain of the virus can be asserted only by more systematic study that is coordinated on a global scale. The data from the Asian countries, and also from most of Africa, where the number of deaths have been relatively low so far, have started showing noticeable modulations only recently, when the fraction of the cases and deaths have become significant. This suggests a possible link between the periodic pattern and the phenomenon of community transmission and infection. If that is the case, then the pattern could be an early indicator of community spreading of the pandemic.

## Data Availability

Only open data used.

https://ourworldindata.org

